# Poor R-Wave Progression and Long-Term Outcomes in the Multi-Ethnic Study of Atherosclerosis (MESA)

**DOI:** 10.1101/2025.08.04.25332992

**Authors:** José Nunes de Alencar, João Vitor Gardelli Trindade, Claudionor Antônio dos Santos Filho, Sandro Pinelli Felicioni, Mariana Fuziy Nogueira De Marchi

**Author notes:** **CORRESPONDING AUTHOR** José Nunes de Alencar Neto, Instituto Dante Pazzanese de Cardiologia, Avenida Dr. Dante Pazzanese No. 500, Vila Mariana, São Paulo 04012-091, Brazil. **Data availability statement:** De-identified MESA data from Exams 1–5 with adjudicated events through calendar year 2015 are available from the NHLBI BioLINCC repository (Accession HLB00640824a) upon approved application and data-use agreements. Access for commercial entities is tiered per participant consent. **Ethics approval:** All participants provided written informed consent and the protocol was approved by the institutional review boards of each participating center. **Clinical trial registration:** ClinicalTrials.gov Identifier NCT00005487. **AI use disclosure:** We used Claude (Anthropic) to (1) assist with R code debugging/structuring (e.g., improving data-cleaning functions, model-reporting tables, and plotting syntax) and (2) provide language editing to improve clarity and flow in American English. All study design, analyses, interpretation, and original scientific content were conceived and written by the authors. The authors verified all code and results and take full responsibility for accuracy. No identifiable patient information was shared with the AI; any material used for editing was de-identified or paraphrased. **Patients or public involvement:** Patients or the public were not involved in the design, or conduct, or reporting, or dissemination plans of our research. **Author Contributions** Conceptualization: José Nunes de Alencar Neto (JNdA) Project Administration: JNdA Writing – Original Draft: JNdA, Sandro Pinelli Felicioni (SPF) and Mariana De Marchi (MDM) Writing – Review & Editing: JNdA, SPF and MDM Guarantor of the Article: JNdA accepts full responsibility for the integrity of the study and the manuscript. All authors had full access to the data, substantially contributed to manuscript revision, and approved the final version.

## Abstract

**Background:** Poor R-wave progression (PRWP) has been interpreted as a marker of anterior myocardial injury since the 1970s, but its temporal stability, substrate, and independent prognostic value in a CAD-free cohort remain uncertain.

**Methods:** We studied 6,323 MESA participants free of clinical cardiovascular disease with standardized 12-lead ECGs at baseline (Exam 1, 2000–02) and after a median of 10 years (Exam 5, 2010–12). PRWP was defined as R-wave amplitude ≤3 mm in V3 with RV3 > RV2. We assessed 10-year test–retest stability (Cohen’s κ), pulmonary/thoracic correlates, mechanistic substrate via cardiac magnetic resonance, high-sensitivity troponin T, NT-proBNP, and polysomnography, and long-term outcomes (all-cause death, cardiovascular death, MACE) over median 14 years using Cox and Fine–Gray models.

**Results:** PRWP prevalence was 3.2% at Exam 1 and 6.2% at Exam 5. Binary test–retest agreement was limited (κ = 0.344; 95% CI 0.283–0.406) despite good continuous R-V3 reliability (ICC = 0.73); only 52% of baseline PRWP persisted at 10 years. Self-reported emphysema (OR 4.48; 2.30–8.22) and current smoking (OR 1.77; 1.14–2.73) were the strongest PRWP predictors. PRWP was not associated with RV volumes, RV ejection fraction, NT-proBNP, or sleep apnea. After adjustment for smoking and emphysema, PRWP was not independently associated with all-cause mortality (HR 1.19; 0.90–1.58), cardiovascular death (HR 1.02; 0.67–1.57), or MACE (HR 0.89; 0.54–1.47). Time-updated and Fine–Gray models confirmed null associations.

**Conclusions:** In a CAD-free multi-ethnic cohort, isolated PRWP is an ECG pattern with limited binary reproducibility, linked to pulmonary exposure and thoracic habitus rather than myocardial substrate, and carries no independent cardiovascular prognostic value.

**Graphical Abstract:** 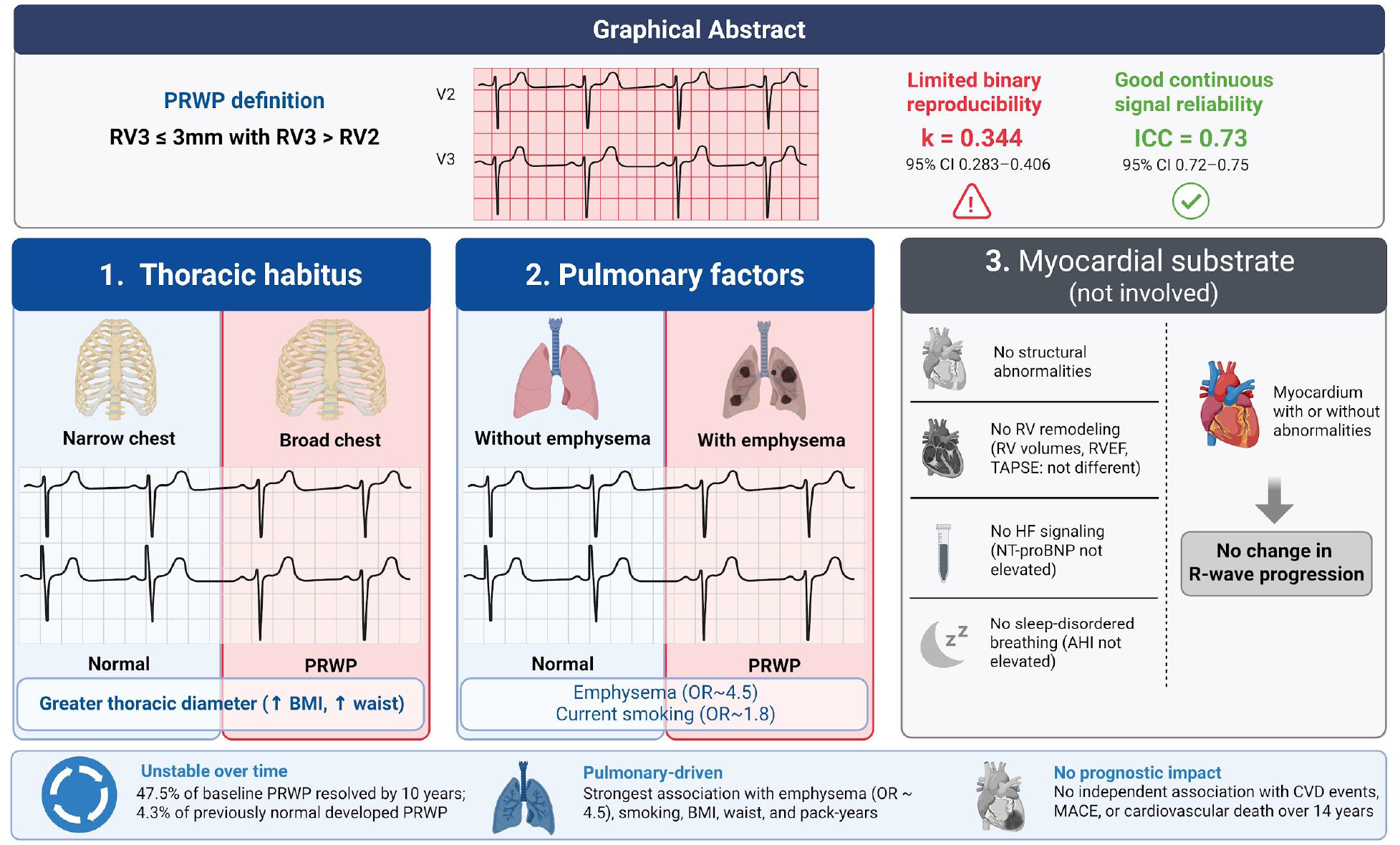

## 1. Introduction

Poor R-wave progression on the resting 12-lead electrocardiogram has been taught for half a century as a marker of prior anterior myocardial injury. The diagnostic tradition arose from small vectorcardiographic case series in the 1970s and 1980s, which enrolled patients with known coronary disease and retrospectively assessed R-wave patterns.^1^ A recent scoping review concluded that the entire evidence base is limited by selection bias, heterogeneous definitions, and the absence of population-based controls.^2^

Two recent prospective reports revived interest in PRWP as a prognostic marker: the Health 2000 survey linked PRWP to two-fold increases in mortality in women,^3^ and the Mini-Finland study linked it to sudden cardiac death.^4^ Both cohorts, however, enrolled participants irrespective of prevalent coronary disease, and their positive signals were concentrated in CAD-positive subgroups.

We recently showed, using baseline CMR late-gadolinium-enhancement imaging in MESA, that isolated PRWP does not identify anterior-wall fibrosis (sensitivity 0–5% against strict CMR criteria).^5^ If PRWP does not reflect scar on gold-standard imaging, what, if anything, does it reflect? Three complementary questions remain unanswered. First, is PRWP on a single baseline ECG a stable patient-level trait, or an incidental electrical configuration that comes and goes? Second, if PRWP has no myocardial scar correlate, does it correspond instead to a pulmonary or thoracic-geometric substrate, as early hyperinflation studies suggested? Third, does isolated PRWP carry independent long-term prognostic weight once CAD is excluded and comorbid substrate is accounted for?

We addressed these three questions in the Multi-Ethnic Study of Atherosclerosis (MESA), a large, contemporary, community-based, multi-ethnic cohort with standardized serial ECGs, CMR-derived right-ventricular function, circulating cardiac biomarkers, and 14 years of adjudicated outcomes.^6^

## 2. Methods

### Study population

MESA is a prospective, NHLBI-funded cohort of 6,814 adults aged 45–84 years, free of clinical cardiovascular disease at enrollment (2000–02), recruited from six U.S. field centers.^6^ The present analysis used baseline ECGs and covariates from Exam 1, repeat ECGs from Exam 5 (2010–12), CMR from Exam 1, biomarkers from the MESA hs-cTnT/NT-proBNP ancillary, and adjudicated events through 31 December 2015. Participants were excluded for Wolff–Parkinson–White pre-excitation, bundle-branch block or major intraventricular conduction delay, pathologic anterior Q-waves by Minnesota coding, or missing key covariates. Institutional review boards at each field center approved the protocol; all participants provided written informed consent.

### ECG acquisition and PRWP definition

Digital 12-lead ECGs were recorded at 25 mm/s and 10 mm/mV (Marquette MAC-PC, GE Healthcare) and transmitted to the MESA ECG Reading Center (Wake Forest), where certified analysts applied Minnesota coding and automated amplitude measurement. PRWP was prespecified as R-wave amplitude ≤3 mm in V3 with RV3 > RV2, applied identically to Exam 1 and Exam 5 ECGs.

### Exposures and covariates

Self-reported emphysema, chronic bronchitis, and asthma were captured at Exam 1 by standardized questionnaire. Smoking status and pack-years were self-reported at Exam 1. Anthropometrics (BMI, waist and hip circumferences) were measured per MESA protocol.

### Mechanistic substrate

Right-ventricular end-diastolic and end-systolic volumes, ejection fraction, mass, and TAPSE were derived from cine CMR at Exam 1 (MESA RV Function ancillary).^7^ High-sensitivity cardiac troponin T and NT-proBNP were measured on stored baseline specimens. Apnea-hypopnea index was measured by in-laboratory polysomnography at Exam 5 in the MESA Sleep ancillary.

### Outcomes

MESA adjudicated events every 9–12 months through centralized review. We analyzed all-cause mortality, MESA Hard-CVD death, and a composite MACE (first of nonfatal myocardial infarction, resuscitated cardiac arrest, or stroke).

### Statistical analysis

For stability, we computed Cohen’s κ with bootstrap 95% CI (1000 replicates) between Exam 1 and Exam 5 PRWP status in participants with eligible paired ECGs. Within-person reliability of continuous R-V3 amplitude was assessed by Pearson correlation and intraclass correlation coefficient (ICC, two-way mixed, absolute agreement). We performed a sensitivity analysis of κ across alternative R-V3 thresholds (≤1.5 mm through ≤5 mm) to evaluate the dependence of binary reproducibility on prevalence (Table S1). Baseline characteristics of the paired versus unpaired cohort were compared to assess survivorship selection (Table S2). We classified each participant into one of four trajectories (stable-normal, incident PRWP, resolved PRWP, persistent PRWP) and compared baseline characteristics across groups. For mechanistic correlates, we fit logistic regression models for PRWP and linear models for continuous R-V3 amplitude, adjusted for age, sex, race/ethnicity, BMI, smoking status, and pack-years; biomarker analyses were log-transformed. For outcomes, we fit baseline Cox models (primary adjustment: age, sex, race, hypertension, diabetes; secondary: + smoking + emphysema), a time-updated Cox model with PRWP status switched at the Exam 5 visit, and a Fine–Gray model for CV death with non-CV death as the competing event. As a sensitivity bound for potential regression-dilution bias, we estimated a disattenuated hazard ratio using κ as the reliability coefficient. Proportional-hazards assumption was assessed via Schoenfeld residuals. All analyses used R 4.5.0.

## 3. Results

### Study population

Of 6,814 enrolled, 6,323 were eligible at Exam 1, and 4,326 had paired eligible ECGs at Exam 5. PRWP was present in 202 at Exam 1 (3.2%) and 288 at Exam 5 (6.2%) (Table 1 and Figure 1).

**Table 1.**
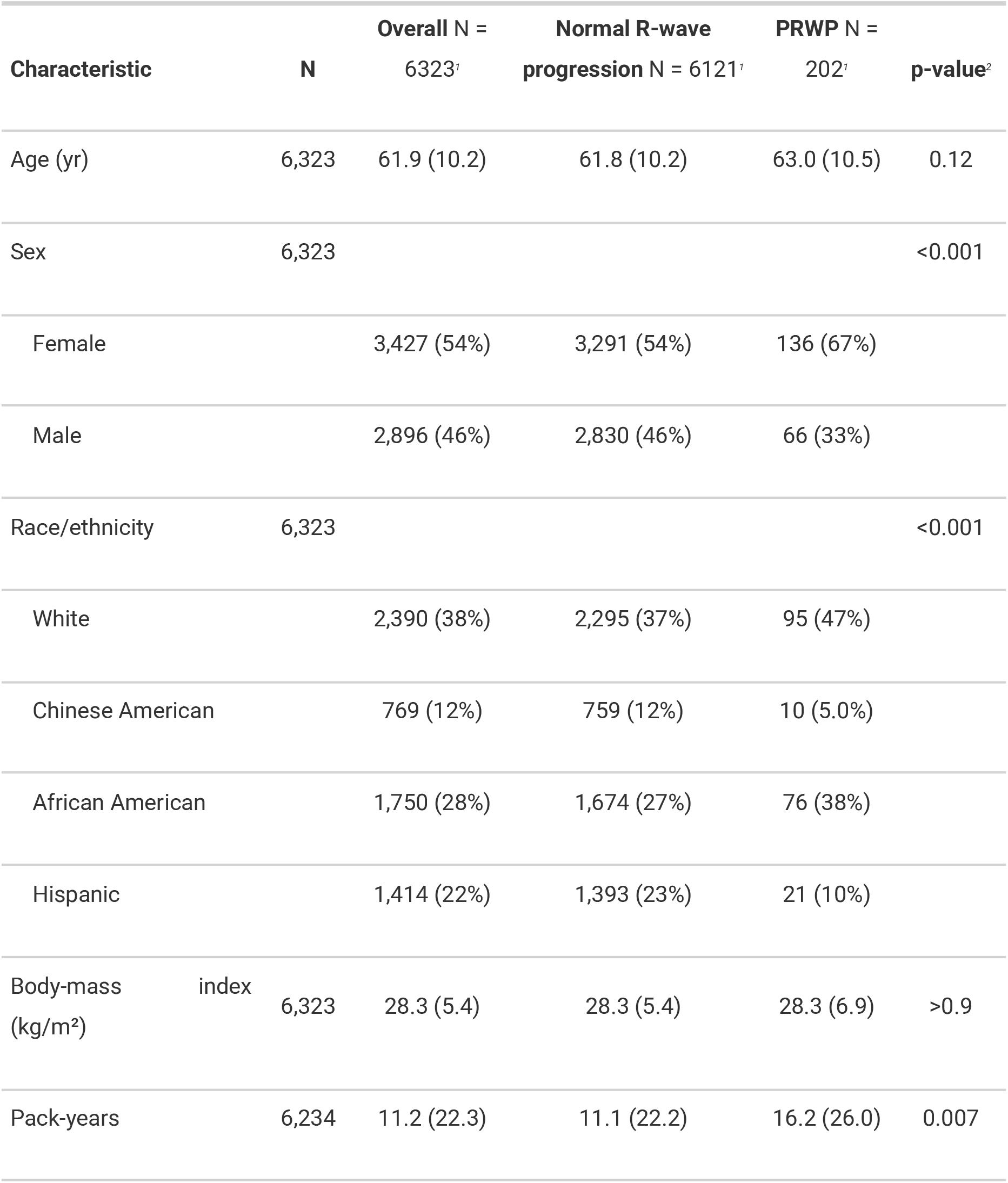
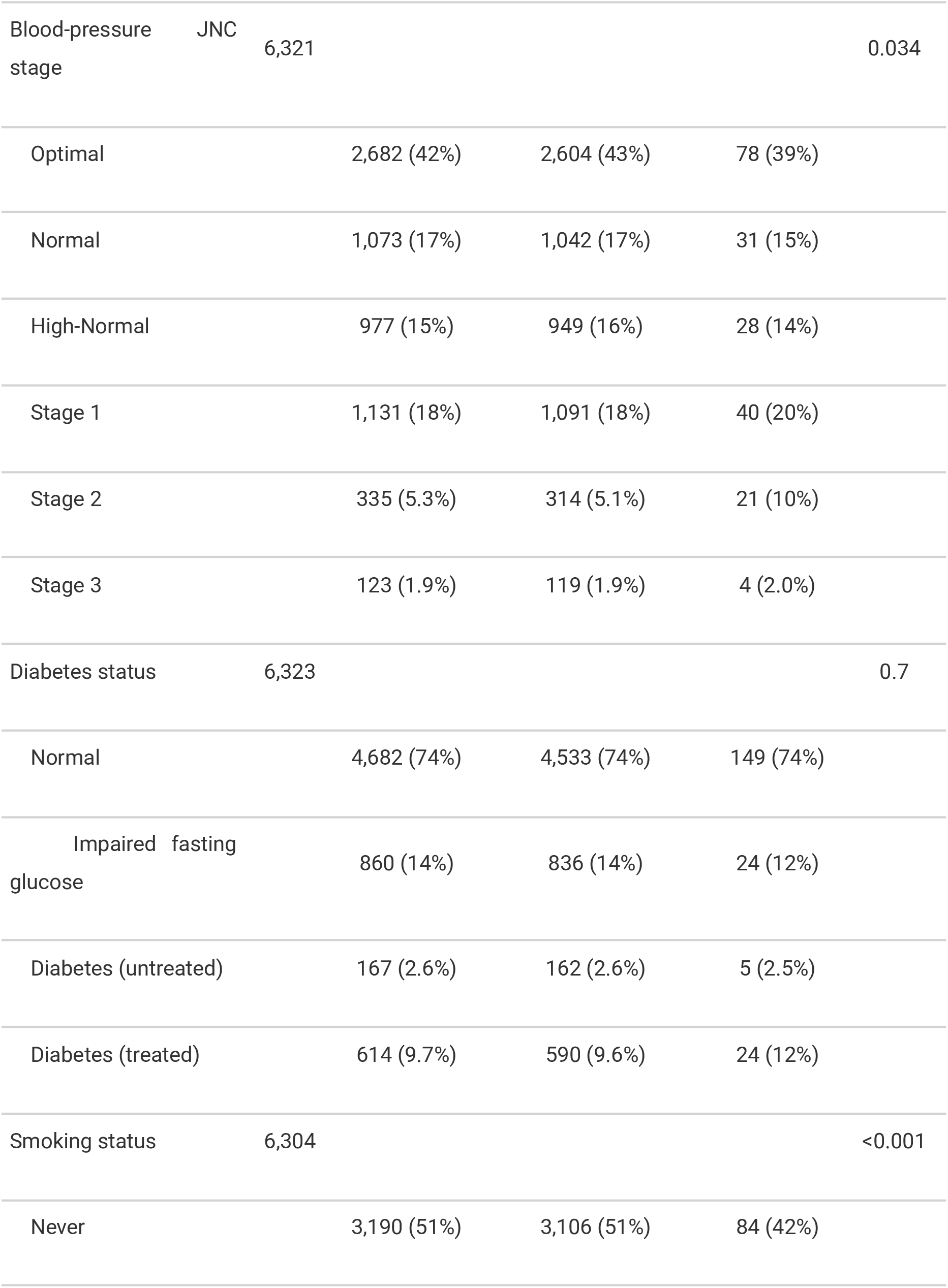
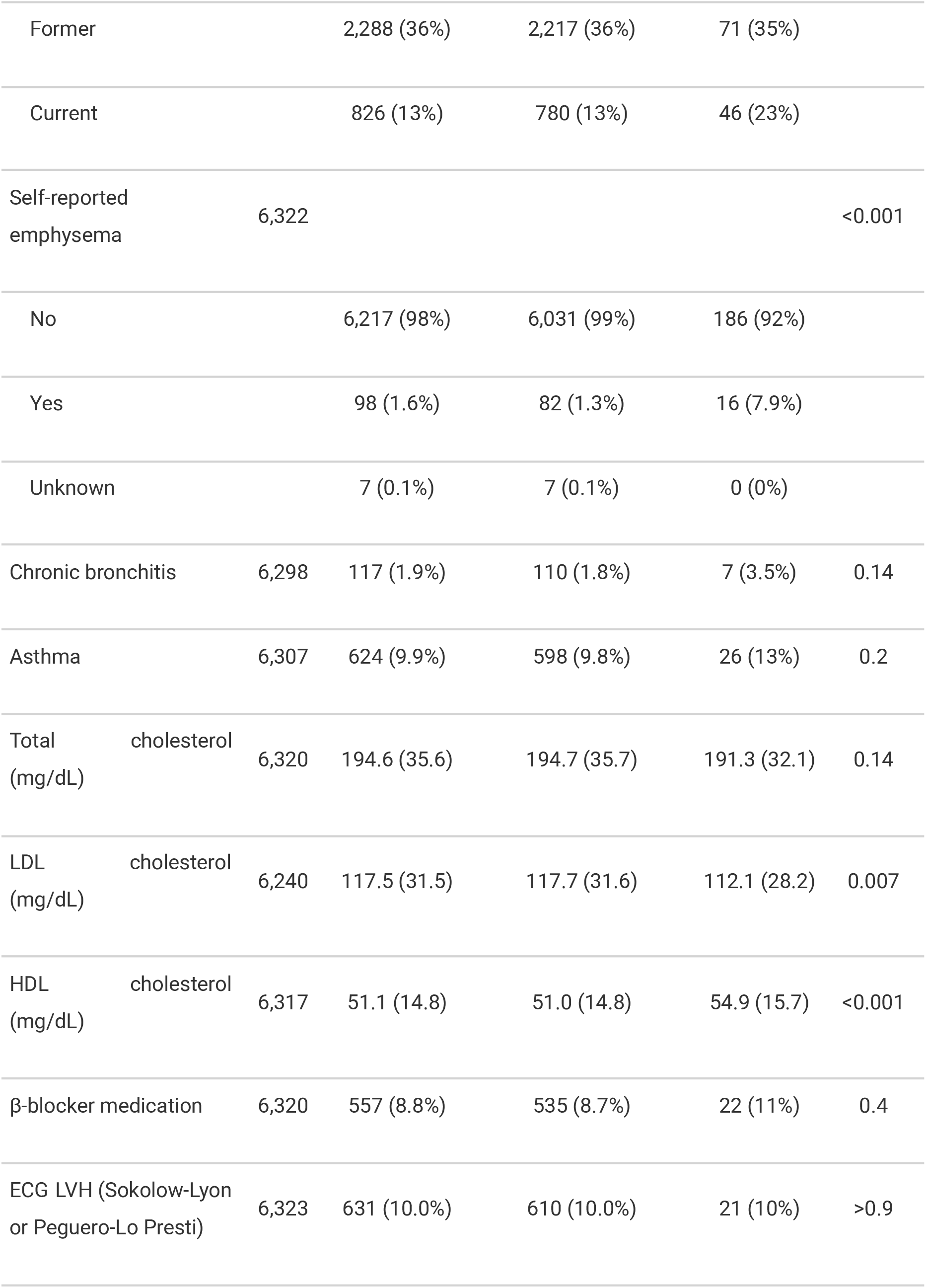
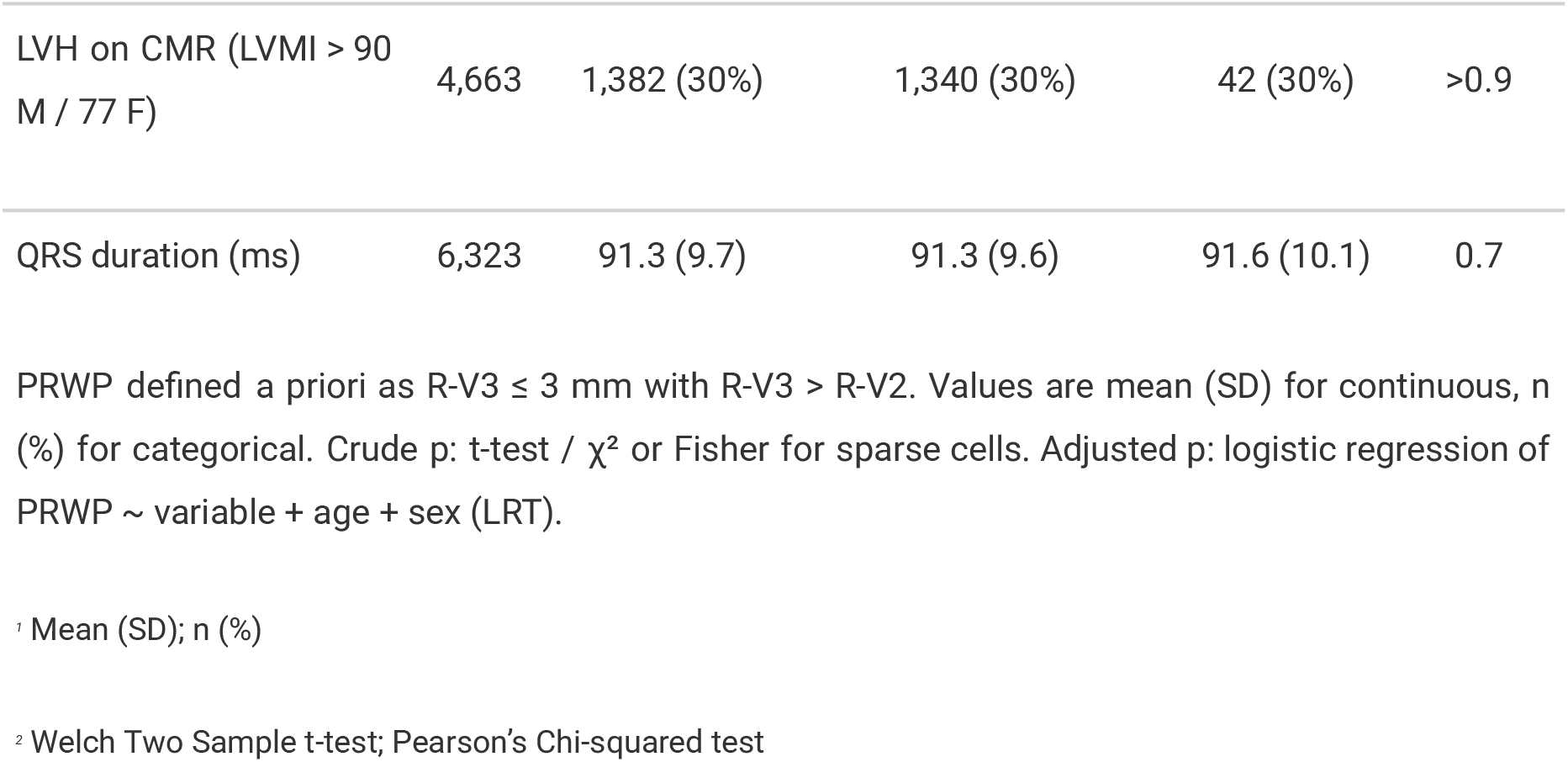
Baseline characteristics at MESA Exam 1, by PRWP status. Analytic cohort n = 6323 (PRWP = 202, 3.2%)

**Figure 1.**
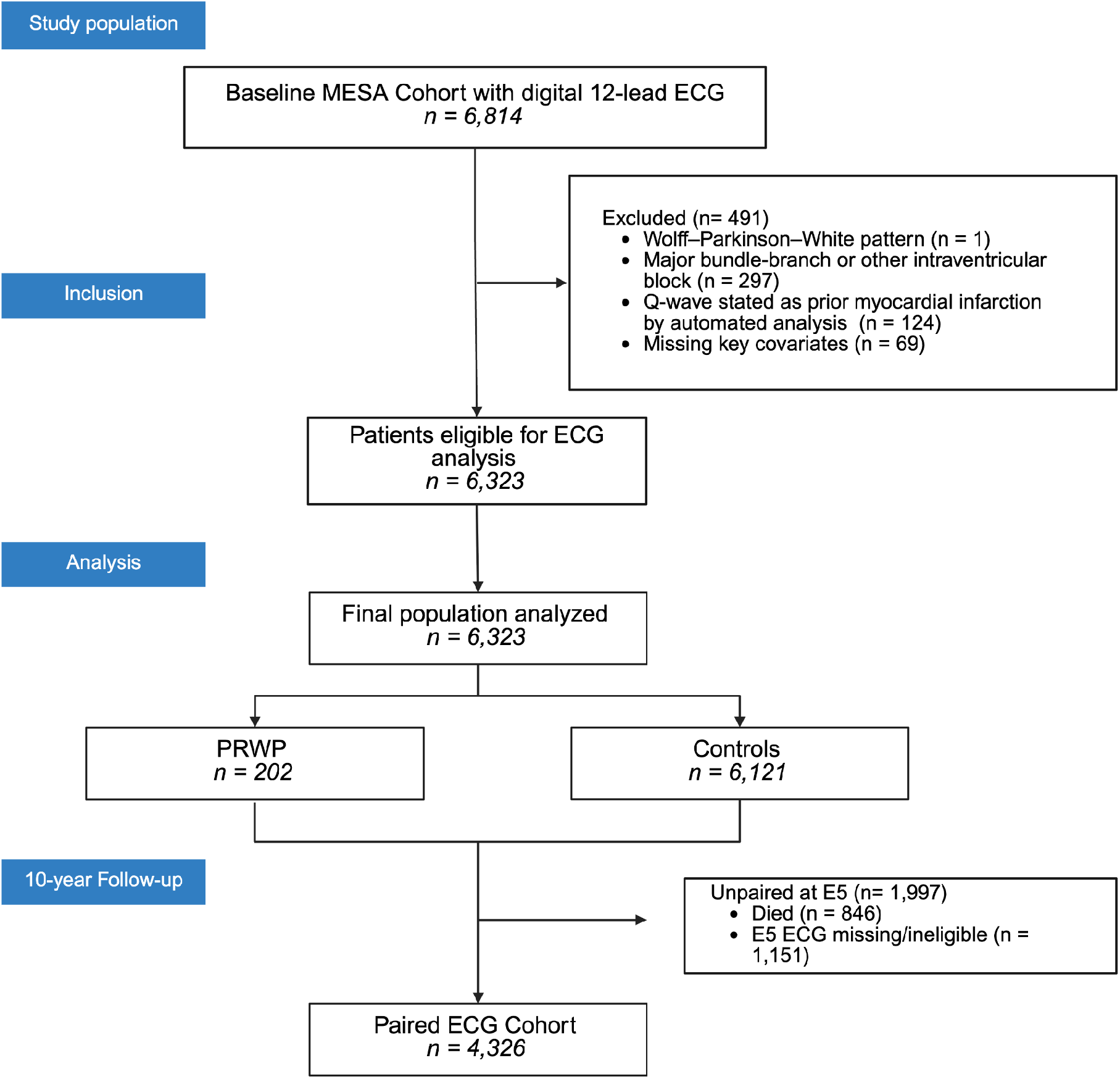
Legend. Study flow diagram.

### Stability

Continuous R-V3 amplitude showed good within-person reliability over a median 9.8 years (ICC = 0.73, 95% CI 0.72–0.75), with a small age-related downward shift (mean Δ −0.51 mm, p < 0.001). When dichotomized at the prespecified threshold, however, binary agreement was only fair (Cohen’s κ = 0.344; 95% CI 0.283–0.406), reflecting the brittleness of a fixed cutpoint near the left tail of the distribution; κ scaled with threshold prevalence from 0.125 at ≤1.5 mm to 0.464 at ≤5 mm (Table S2).

Among 4,326 participants with paired ECGs (Figure 2), roughly half of baseline PRWP resolved by Exam 5 (67/141, 47.5%), while 182 previously normal participants (4.3% of 4,185) developed new PRWP — accounting for the rise in prevalence from 3.2% to 6.2%. Persistent and resolved PRWP groups were enriched for female sex (70%) and self-reported emphysema (5.4–6.0% vs 0.9% in stable-normal); persistent PRWP had the highest current-smoker prevalence (23%).

**Figure 2.**
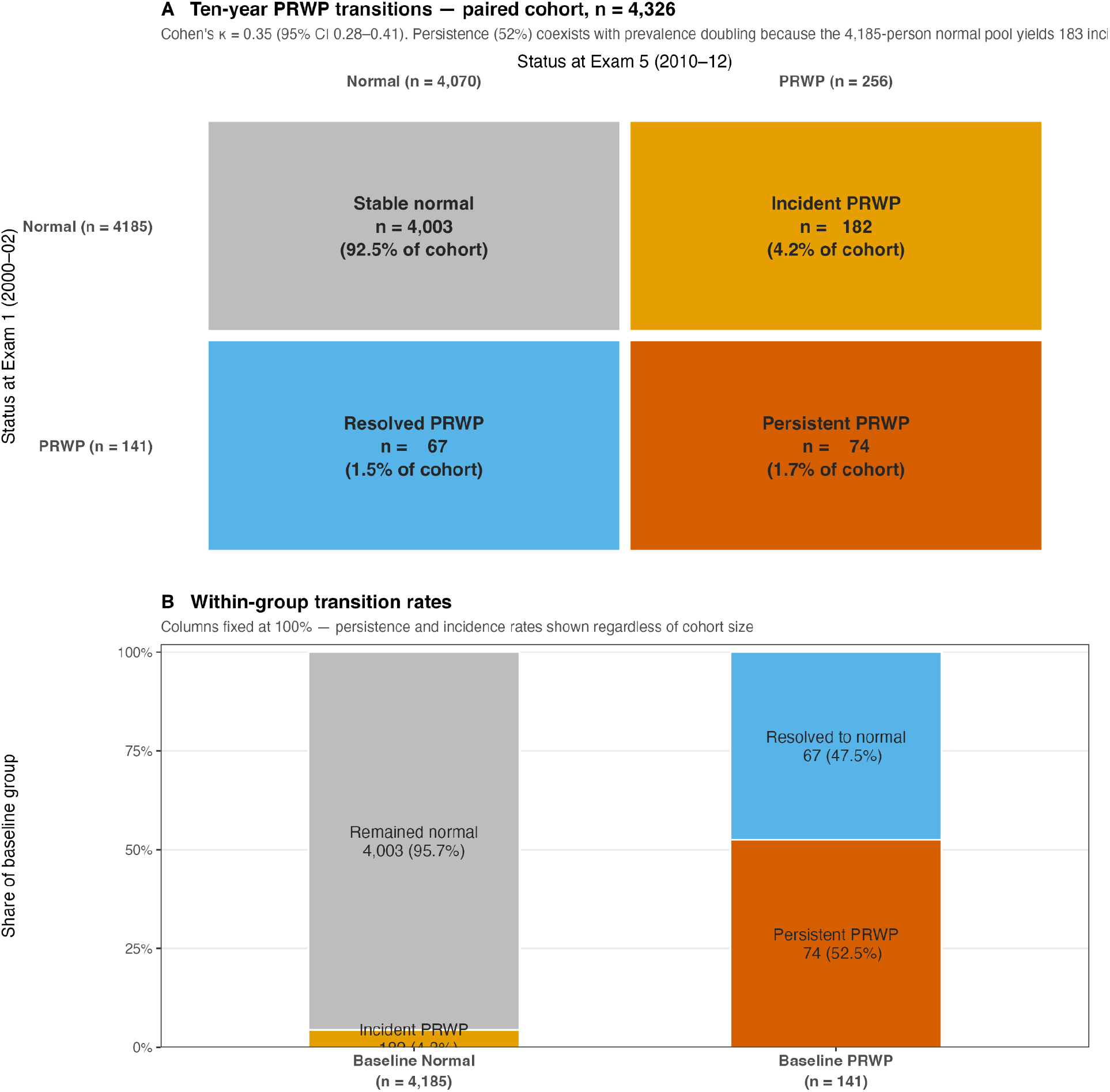
Legend. Ten-year PRWP transitions in the paired ECG cohort (n = 4,326). (A) Mosaic plot showing the cross-classification of PRWP status at Exam 1 (2000–02) versus Exam 5 (2010–12). Cell area is proportional to group size. Of 141 baseline PRWP participants, 74 (52.5%) persisted and 67 (47.5%) resolved; of 4,185 baseline-normal participants, 182 (4.3%) developed incident PRWP. Cohen’s κ = 0.35 (95% CI 0.28–0.41). (B) Stacked bar chart showing within-group transition rates, with columns normalized to 100% of each baseline group regardless of absolute size. PRWP was defined as R-wave amplitude ≤3 mm in V3 with RV3 > RV2.

The paired cohort had substantially lower follow-up mortality than the unpaired cohort (6.8% vs 42.7%; Table S2), indicating survivorship selection; however, baseline PRWP prevalence was equivalent in both groups (3.25% vs 3.04%), arguing against differential attrition by PRWP status.

### Pulmonary and thoracic correlates

In a logistic model including age, sex, race/ethnicity, BMI, waist, smoking, pack-years, emphysema, and chronic bronchitis, self-reported emphysema was the strongest predictor of baseline PRWP (OR 4.48; 95% CI 2.30–8.22; p=3.4×10^−6^). Current smoking conferred OR 1.77 (1.14–2.73; p=0.010). In adjusted linear models of continuous R-V3 amplitude, emphysema was associated with a −1.95 mm difference (95% CI −2.96 to −0.93); each 1 kg/m^2^ increment of BMI was associated with −0.056 mm (−0.081 to −0.031); each 1 cm of waist circumference, −0.013 mm (−0.022 to −0.004); and each pack-year, −0.010 mm (−0.016 to −0.004). All effects persisted after joint adjustment.

### Mechanistic substrate

PRWP was not associated with CMR-measured RV end-diastolic volume (adjusted β +1.2 mL; p=0.60), RV end-systolic volume (+0.5 mL; p=0.64), RV ejection fraction (−0.1%; p=0.86), or TAPSE (−0.9 cm; p=0.23); RV mass was borderline (+0.65 g; p=0.053). NT-proBNP was not elevated (log β 0.13; p=0.40). Apnea-hypopnea index at Exam 5 was not elevated (β −0.03; p=0.42). High-sensitivity cardiac troponin T was modestly but significantly higher in PRWP (adjusted log β 0.12; 95% CI 0.05–0.19; p=0.0009), corresponding to absolute geometric means of 7.2 ng/L in PRWP vs 6.3 ng/L in controls — both well below the 14 ng/L 99th-percentile reference value. Taken together, the mechanistic pattern is characterized by thoracic voltage attenuation (chest-wall habitus, parenchymal emphysema, cumulative smoking) without detectable right-heart remodeling, heart-failure signaling, or sleep-disordered breathing.

### Long-term outcomes

Over a median follow-up of 14.1 years (IQR 13.5–14.7), 1,141 participants died, 607 experienced hard cardiovascular events (defined as cardiovascular death, nonfatal myocardial infarction, stroke, or heart failure), and 517 experienced MACE. In the prespecified baseline Cox model (age, sex, race, hypertension, diabetes), PRWP was borderline-associated with all-cause mortality (HR 1.31; 95% CI 0.99–1.73; p=0.059). Additional adjustment for current smoking and self-reported emphysema attenuated the effect to HR 1.19 (0.90–1.58; p=0.23). PRWP was not associated with hard CVD events (HR 1.02; 0.67–1.57; p=0.915) or MACE (HR 0.89; 0.54–1.47) after the same adjustment. A time-updated Cox model, which reclassified PRWP status at the Exam 5 visit, yielded HR 0.91 (0.58–1.41; p=0.67) for all-cause mortality (Figure 3). A Fine–Gray subdistribution-hazards model for cardiovascular death, with non-cardiovascular death as the competing event, yielded PRWP subHR 0.94 (0.60–1.48; p=0.80).

**Figure 3.**
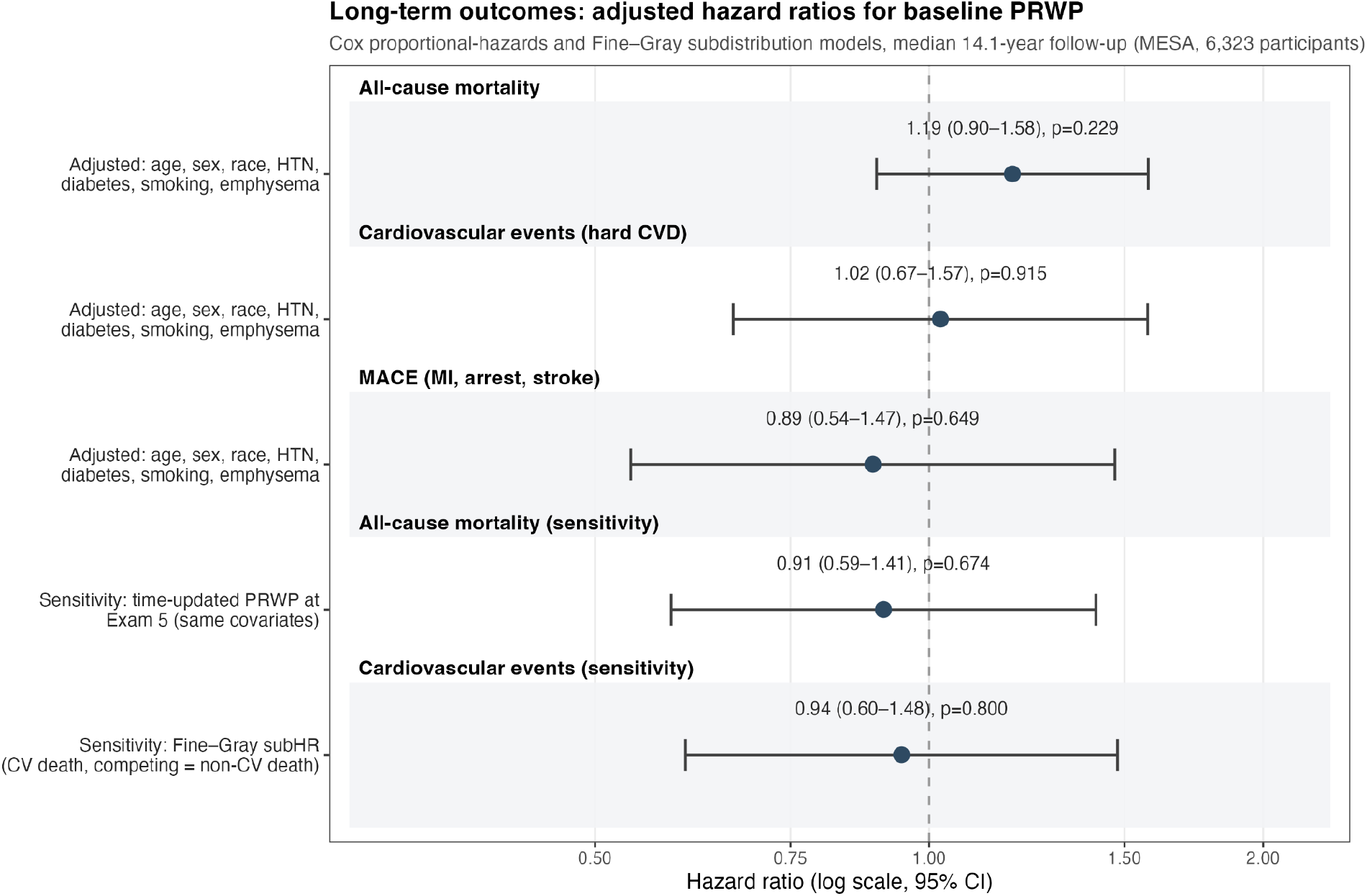
Legend. Adjusted hazard ratios for baseline PRWP and long-term outcomes over median 14.1 years (n = 6,323). The top three rows show Cox proportional-hazards models adjusted for age, sex, race/ethnicity, hypertension, diabetes, smoking, and emphysema. Sensitivity analyses include a time-updated Cox model reclassifying PRWP at Exam 5 and a Fine–Gray subdistribution-hazards model for cardiovascular death with non-cardiovascular death as the competing event. All confidence intervals cross 1.0. PRWP is defined as R-wave amplitude ≤3 mm in V3 with RV3 > RV2.

## 4. Discussion

In this CAD-free, multi-ethnic community cohort, we found three things. The binary PRWP flag had poor test–retest agreement over 10 years (κ = 0.34), even though the continuous R-V3 amplitude was reasonably reproducible (ICC = 0.73) — a gap explained by the sensitivity of a fixed cutpoint applied near the tail of the distribution. PRWP tracked pulmonary and thoracic factors (emphysema, OR 4.48; smoking; BMI; waist circumference; pack-years) but showed no association with RV remodeling, NT-proBNP, or sleep-disordered breathing. And PRWP carried no independent association with cardiovascular death or MACE over 14 years; the borderline signal for all-cause mortality (HR 1.31) attenuated to 1.19 after adjustment for smoking and emphysema, and disappeared entirely in time-updated and competing-risks models.

No single one of these results is definitive. Taken together, they point in the same direction: PRWP in the absence of CAD is a voltage pattern shaped by the chest wall and lungs, not by the myocardium. We previously showed that isolated PRWP fails to identify anterior fibrosis on CMR-LGE in MESA (sensitivity 0–5%).^5^ The present data add that the pattern is temporally unstable and prognostically inert once its pulmonary substrate is accounted for. The modest troponin elevation in PRWP participants (geometric means 7.2 vs 6.3 ng/L, both well below the 14 ng/L clinical threshold) is the one finding that does not fit this interpretation cleanly and deserves mention, but it was not accompanied by any structural, neurohormonal, or outcome correlate.

The positive associations reported from the Finnish Health 2000 and Mini-Finland cohorts are best explained by case-mix: both studies included participants with prevalent CAD, and their mortality signals concentrated in CAD-positive subgroups.^3,4^ MESA excluded clinical CAD at enrollment, and no signal persisted.

### Clinical implications

Automated commercial ECG reports routinely flag isolated PRWP as “abnormal,” prompting downstream echocardiography and stress testing. Our findings, combined with the CMR diagnostic null, argue that in patients without established coronary disease, isolated PRWP should not drive cardiac workup; when clinically relevant, its association with emphysema, smoking, and thoracic habitus suggests pulmonary evaluation may be more productive.

### Limitations

Emphysema was self-reported, not CT-quantified; CT-based metrics from the MESA-Lung ancillary were not available in the BioLINCC release and represent a natural follow-up. Only two serial ECGs were analyzed; additional time points would refine incidence/reversion estimates. The rise in PRWP prevalence from 3.2% to 6.2% between exams, consistent with age-related chest-geometry changes, means our stability analysis is conservative regarding individual-level reproducibility. Stability analyses were restricted to survivors with a usable Exam-5 ECG; however, baseline PRWP prevalence was equivalent in paired and unpaired cohorts (3.25% vs 3.04%; Table S2), arguing against differential attrition by PRWP status. Finally, if the limited binary reproducibility of PRWP reflects measurement error, the baseline Cox HR of 1.31 is attenuated toward the null; a regression-dilution–corrected estimate using κ = 0.34 yields a disattenuated HR of approximately 2.2, which should be interpreted as a sensitivity bound rather than a point estimate.

## 5. Conclusions

Isolated poor R-wave progression, evaluated in a large multi-ethnic cohort without clinical cardiovascular disease, is a, ECG pattern with limited binary reproducibility over time, associated with pulmonary exposure and thoracic habitus rather than with imaging-detectable myocardial substrate, and carries no independent long-term cardiovascular prognostic weight. These findings support treating isolated PRWP as an electrical configuration of thoracic origin rather than a cardiac alarm.

## Data Availability

De-identified MESA data from Exams 1-5 with adjudicated events through calendar year 2015 are available from the NHLBI BioLINCC repository (Accession HLB00640824a) upon approved application and data-use agreements. Access for commercial entities is tiered per participant consent.

https://biolincc.nhlbi.nih.gov/studies/mesa/

## Results

Baseline characteristics of the analytic cohort at MESA Exam 1 (2000–02), stratified by PRWP status. PRWP was defined a priori as R-wave amplitude ≤3 mm in V3 with RV3 > RV2. Continuous variables are presented as mean (SD) and categorical variables as n (%). P-values are from Welch two-sample t-tests for continuous variables and Pearson’s chi-squared test (or Fisher’s exact test for sparse cells) for categorical variables. LVH on CMR was available in 4,663 participants with adequate cine imaging. Abbreviations: BMI, body-mass index; CMR, cardiac magnetic resonance; JNC, Joint National Committee; LDL, low-density lipoprotein; HDL, high-density lipoprotein; LVH, left ventricular hypertrophy; LVMI, left ventricular mass index.

## Notes

**Conflicts of Interest Disclosure:** There is no conflict of interest to disclose.

### Competing Interest Statement

The authors have declared no competing interest.

### Clinical Trial

NCT00005487

### Funding Statement

The Multi-Ethnic Study of Atherosclerosis (MESA) is supported by contracts from the National Heart, Lung, and Blood Institute (NHLBI), National Institutes of Health. The content is solely the responsibility of the authors and does not necessarily represent the official views of the NHLBI or the National Institutes of Health.

### Author Declarations

All participants provided written informed consent and the protocol was approved by the institutional review boards of each participating center.

### Summary of Updates

This version of the manuscript has been revised to include one additional figure and two new authors. Author affiliations have been updated accordingly.

